# Fetal Growth Disorders Detection During First Trimester Gestation Through Comprehensive Maternal Circulating DNA Profiling

**DOI:** 10.1101/2024.12.21.24319487

**Authors:** Rene Cortese, Kylie Cataldo, Justin Hummel, Gracie Smith, Madison Ortega, Madison Richey, Hung Winn, David Gozal, Jean Ricci Goodman

**Affiliations:** Department of Gynecology, Obstetrics and Women’s Health. University of Missouri. Columbia, MO, 65212, USA; Department of Pediatrics. School of Medicine. University of Missouri. Columbia, MO, 65212, USA; Institute for Data Science and Informatics, University of Missouri, Columbia, MO, 65212, USA; Present address: Saint Luke’s Hospital Maternal Fetal Medicine Specialists. Kansas City, MO, 64111, USA; Present address: Joan C. Edwards School of Medicine. Marshall University, Huntington, WV 25755, USA

## Abstract

**BACKGROUND:** Early diagnosis, close follow-up and timely delivery constitute the main elements for appropriate detection and management of Fetal Growth Disorders (FGD). We hypothesized that fetoplacental FGD-associated alterations can be detected in circulating DNA (cirDNA) samples isolated from maternal blood, as early as the first gestational trimester. To study whether markers in maternal cirDNA may facilitate FGD early detection, we profiled plasma cirDNA from maternal samples prospectively collected during first gestational trimester.

**METHODS:** Plasma cirDNA was isolated from samples prospectively collected during first trimester gestation (n=56). Small, Large and Appropriate for Gestational Age (SGA n=11, LGA n=18, and AGA n=29, respectively) status was determined at birth according to weight and gestational age. cirDNA amount, fragmentation, mitochondrial/nuclear ratio and cirDNA methylation profiles were quantified using qPCR-based assays. Machine learning approaches were applied to build a molecular signature for prediction of LGA and SGA. Prediction accuracy was assessed by Receiving-Operating Curve (ROC) analysis, and Positive and Negative Predictive values (PPV and NPV, respectively) were calculated.

**RESULTS:** Total concentration of plasma cirDNA, cirDNA fragmentation and ratio of mitochondrial/nuclear cirDNA were increased in SGA and LGA compared to AGA pregnancies. DNA methylation profiles also shown distinctive patterns. Out of the 10 selected loci, we detected 5 genes (*HSD2*, *RASSF1*, *CYP19A1*, *IL10*, and *LEP*) showing significant differential methylation differences (p<0.05) across the SGA, AGA and LGA samples at first trimester. We combined these molecular and epigenetic cirDNA markers in a signature that reliably discriminates between FGD and AGA pregnancies with high accuracy (AUC>0.95), achieving 88.8% PPV and 85.7% NPV.

**CONCLUSIONS:** Our findings show that maternal blood cirDNA profiles accurately detects early gestation FGD. The proposed novel marker panel hold great potential for implementation of low invasive approaches for reliable prediction of FGDs, enabling a disruptive path toward precision medicine in FGD.

## MAIN TEXT INTRODUCTION

According to the United Nations, nearly 400,000 babies are born daily worldwide. Of those, it is estimated that 8.6% are born small for gestational age (SGA, <10th percentile), 80.9% appropriate for gestational age (AGA, 10-90th percentile), and 10.5% large for gestational age (LGA, >90th percentile)(1). Many potential adverse health outcomes that could affect the mother and fetus are associated with fetal growth disorders (FGD) during pregnancy (2, 3).

Notwithstanding, we are unaware of any intervention that is currently available and can improve outcomes in SGA and LGA pregnancies targeting the affected molecular and physiological pathways. Hence, early diagnosis, close monitoring and timely delivery constitute the main elements for the appropriate detection and management of pregnancies with FGD(4). Nowadays, screening of fetal growth abnormalities is based on fetal biometric measurements using ultrasound in which serial sonographic assessments of fetal size over time are used to estimate fetal growth and identify any deviation from normative trajectories. However, besides the unprecedented technological development in the field, errors and approximations still hinder FGD detection and assessment(5). Hence, there is an unmet need for reliable early pre-natal biomarkers of fetal growth that can be assessed using low-invasive approaches.

FGDs are associated with fetal (6), placental (7), and maternal anomalies (8). However, whether those anomalies have different weight in FGD etiology throughout the extent of pregnancy remains to be determined. Hence, it is reasonable that an analyte that will reflect such complexity, such as circulating DNA (cirDNA) in maternal blood may hold the potential for sensitive and specific assessment of FGDs at different gestational stages, paving the way for the development of molecular diagnostic tools and providing insights on the temporal differences in fetoplacental and maternal pathophysiology in FGD.

Since the initial report by Dennis Lo and cols (9), numerous studies have been conducted to determine the origin and dynamics of the fetal cirDNA in maternal plasma (reviewed in (10)). cirDNA in plasma derives from the fetus, the placenta and the mother (11), and the relative contribution of each source to the total amount of DNA recovered from maternal plasma or serum is highly dynamic and followed by rapid clearance after birth (12). Thus, the assessment of cirDNA in maternal blood can provide valuable evidence for fetoplacental health and disease throughout pregnancy (10) .

In this study, we hypothesized that fetoplacental FGD-associated alterations can be detected in cirDNA samples isolated from maternal blood. As a corollary, we propose that cirDNA markers can predict the occurrence of SGA and LGA pregnancies, as early as in first trimester of gestation, when assessed either individually, or in combination in a marker panel generated via machine learning approaches.

## RESULTS

### Molecular characterization of plasma cirDNA in first trimester SGA, LGA and AGA pregnancies

The total concentration of plasma cirDNA in SGA (mean cirDNA=9.40 ± 5.49 ng/mL) and LGA (mean cirDNA=12.36 ± 4.24 ng/mL) was not significantly higher than in AGA pregnancies (mean cirDNA=8.48 ± 3.16 ng/mL) (p=0.281; Kruskal-Wallis test) (Figure 2A). The fraction of fragmented DNA was significantly increased in LGA (mean DFI=2.78 ± 1.46) and in SGA (mean DFI=2.37 ± 0.45) compared to AGA pregnancies (mean DFI=1.56 ± 0.71) (p=0.001; Kruskal-Wallis test; p_LGA-AGA_=0.004, and p_SGA-AGA_=0.019, Dunn’s test) (Figure 2B). Noteworthy, we also detected significant differences (p=0.005, Kruskal-Wallis test) in the ratio of mitochondrial/nuclear DNA among SGA, LGA and AGA pregnancies (Figure 2C). Compared with AGA pregnancies, p*ost hoc* analysis revealed a significant increase in LGA (mean MNR=1.20 ± 1.18 and 0.23 ± 0.22, for LGA and AGA pregnancies, respectively, p=0.004, Dunn’s test), and a non-significant increase in SGA (mean MNR=0.78 ± 1.02 and 0.23 ± 0.22, for SGA and AGA pregnancies, respectively, p=0.274, Dunn’s test).

**Figure 1:**
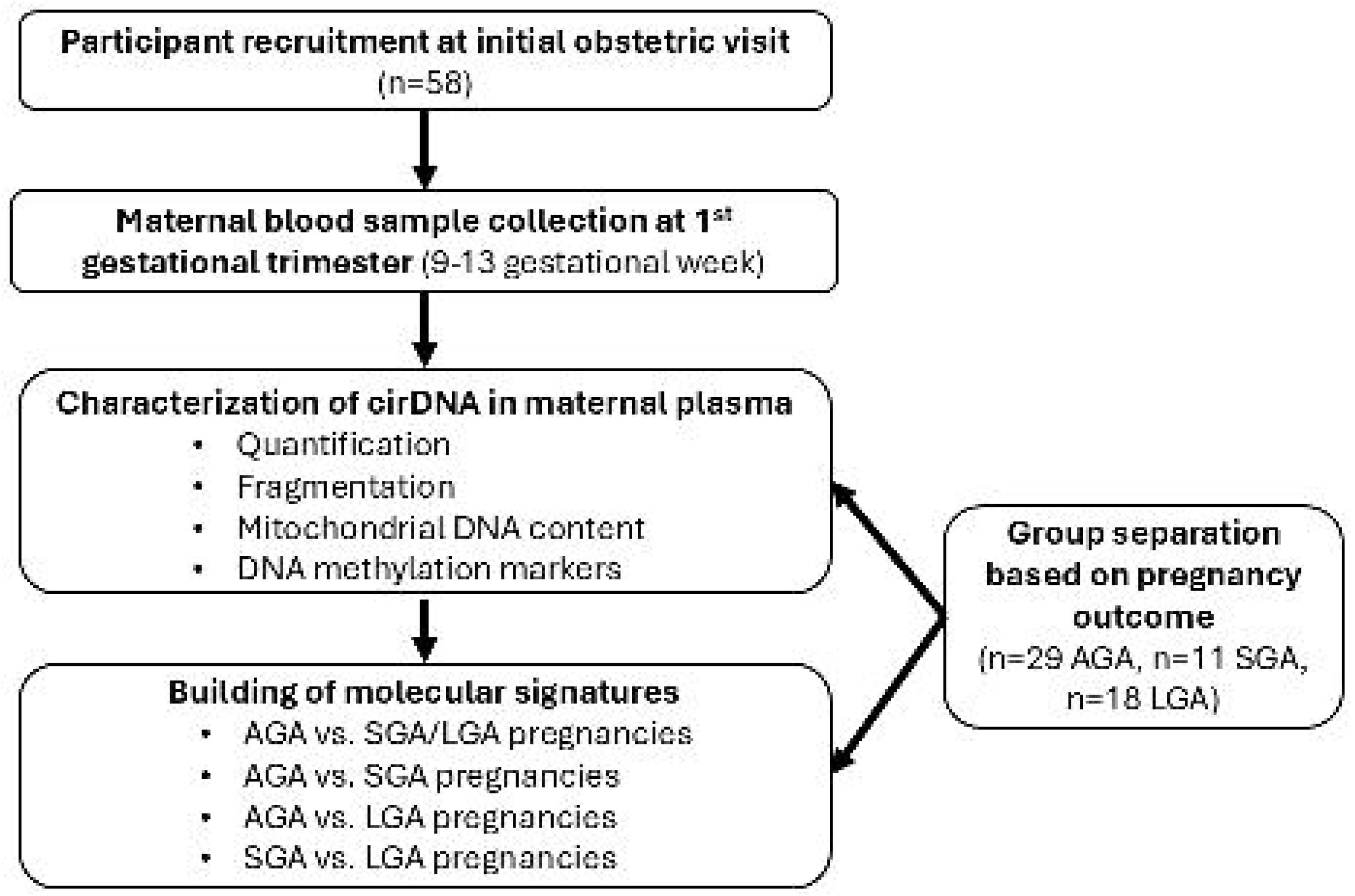
Study design. Participants were prospectively recruited at their initial Obstetrics visit and first gestational trimester (gestational week 9-13) maternal blood samples were collected. Plasma cirDNA was isolated and characterized by assessing quantity, fragmentation, mitochondrial DNA content and DNA methylation status in 10 candidate genes using qPCR- based methods. Participants were grouped based on the outcome of the pregnancy taking in account gestational age and weight at delivery, following the revised reference chart for the US(27), into three groups SGA (< 10^th^ percentile, n=11), LGA (>90^th^ percentile, n=18) and AGA (10^th^-90^th^ percentile, n=29). Plasma cirDNA markers were evaluated individually and combined in molecular signatures differentiating among AGA, SGA, and LGA pregnancies.

**Figure 2:**
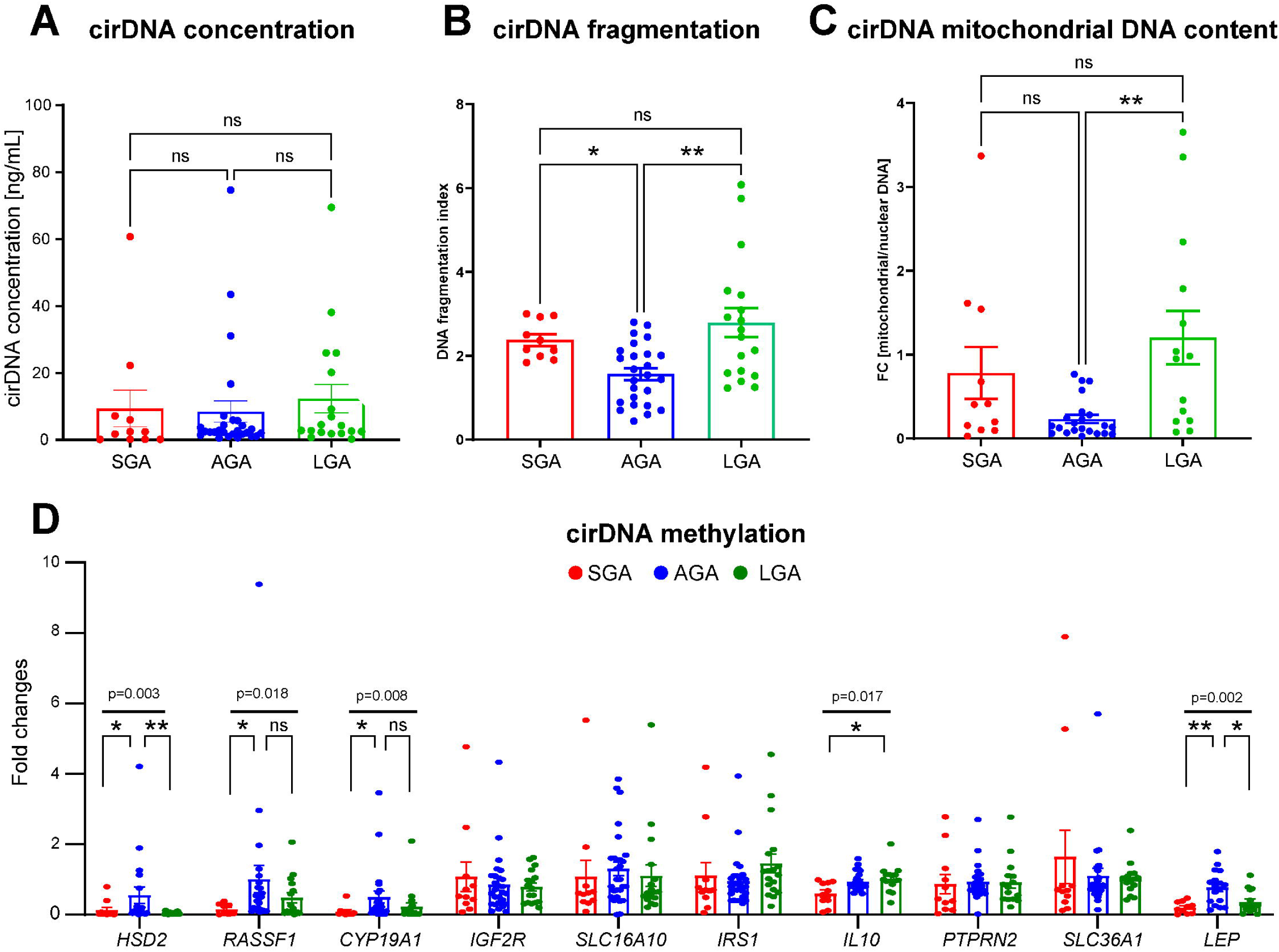
cirDNA profiling in SGA, LGA and AGA pregnancies. A) Total plasma cirDNA concentration was increased in SGA and LGA compared with AGA pregnancies (mean cirDNA=9.40 ± 5.49 ng/mL, 12.36 ± 4.24 ng/mL, and 8.48 ± 3.16 ng/mL, for SGA, LGA and AGA respectively), although the differences were not statistically significant (p=0.281). B) Fraction of fragmented (apoptotic) cirDNA was significantly increased (p=0.001) in SGA and LGA compared with AGA pregnancies (mean DFI= DFI=2.37 ± 0.45, 2.78 ± 1.46, and 1.56 ± 0.71, for SGA, LGA and AGA respectively). C) Ratio of mitochondrial/nuclear DNA was significantly different (p=0.005) in SGA and LGA compared with AGA pregnancies (mean MNR= 0.78 ± 1.02, 1.20 ± 1.18 and 0.23 ± 0.22, for SGA, LGA and AGA pregnancies, respectively). D) Differential cirDNA methylation in 10 candidate genes We observed significant differences among SGA, LGA and AGA pregnancies in 5 out of the 10 genes (p_HSD2_=0.003, p_RASSF1_=0.018, p_CYP19A1_=0.008, p_IL10_=0.017 and, p_LEP_=0.002; Kruskal-Wallis test). SGA, AGA and LGA groups are represented by red, blue and green bars, respectively. p-values correspond to Kruskal-Wallis test. Significance of *ad hoc* comparisons was calculated using Dunn’s test and are expressed as **: p<0.05, *: p<0.01, n.s.: non-significant.

Next, we investigated cirDNA methylation of the 10 selected candidate genes (13–22) (Figure 2D). Significant differences in 5 out of the 10 genes were present (p_HSD2_=0.003, p_RASSF1_=0.018, p_CYP19A1_=0.008, p_IL10_=0.017 and, p_LEP_=0.002; Kruskal-Wallis test). *Post hoc* analysis revealed significant differences between SGA and AGA in 4 genes (p_HSD2_=0.034, p_RASSF1_=0.0182, p_CYP19A1_=0.013, and p_LEP_=0.004, Dunn’s test), between LGA and AGA in 2 genes (p_HSD2_=0.005, and p_LEP_=0.028, Dunn’s test), and between SGA and LGA in the *IL10* gene (p=0.013, Dunn’s test).

### Multivariate analysis of plasma cirDNA profiles in first trimester SGA, LGA and AGA pregnancies

Principal Component Analysis (PCA; Figure 3) showed that samples corresponding to SGA, LGA and AGA pregnancies clustered separately based upon the variables assessed in cirDNA isolated from first trimester maternal blood (Figure 3A). As shown in Figure 3B, decrease in cirDNA methylation in *IL10, SLC36A1, PTPRN2* and *LEP* underlie sample distribution shifts towards the SGA samples. Conversely, decreased cirDNA methylation in the *HSD2* gene and increased DNA fragmentation dragged sample distribution towards LGA samples. Correlation with clinical variables also demonstrated a positive correlation between the levels of mitochondrial DNA in plasma and the presence of maternal hypertension. Remarkably, cirDNA methylation in *SLC16A10*, *IRS1*, *PTPRN2* and *SLC36A1* was positively correlated with maternal age, but not with other clinical variables (Figure 3C).

**Figure 3:**
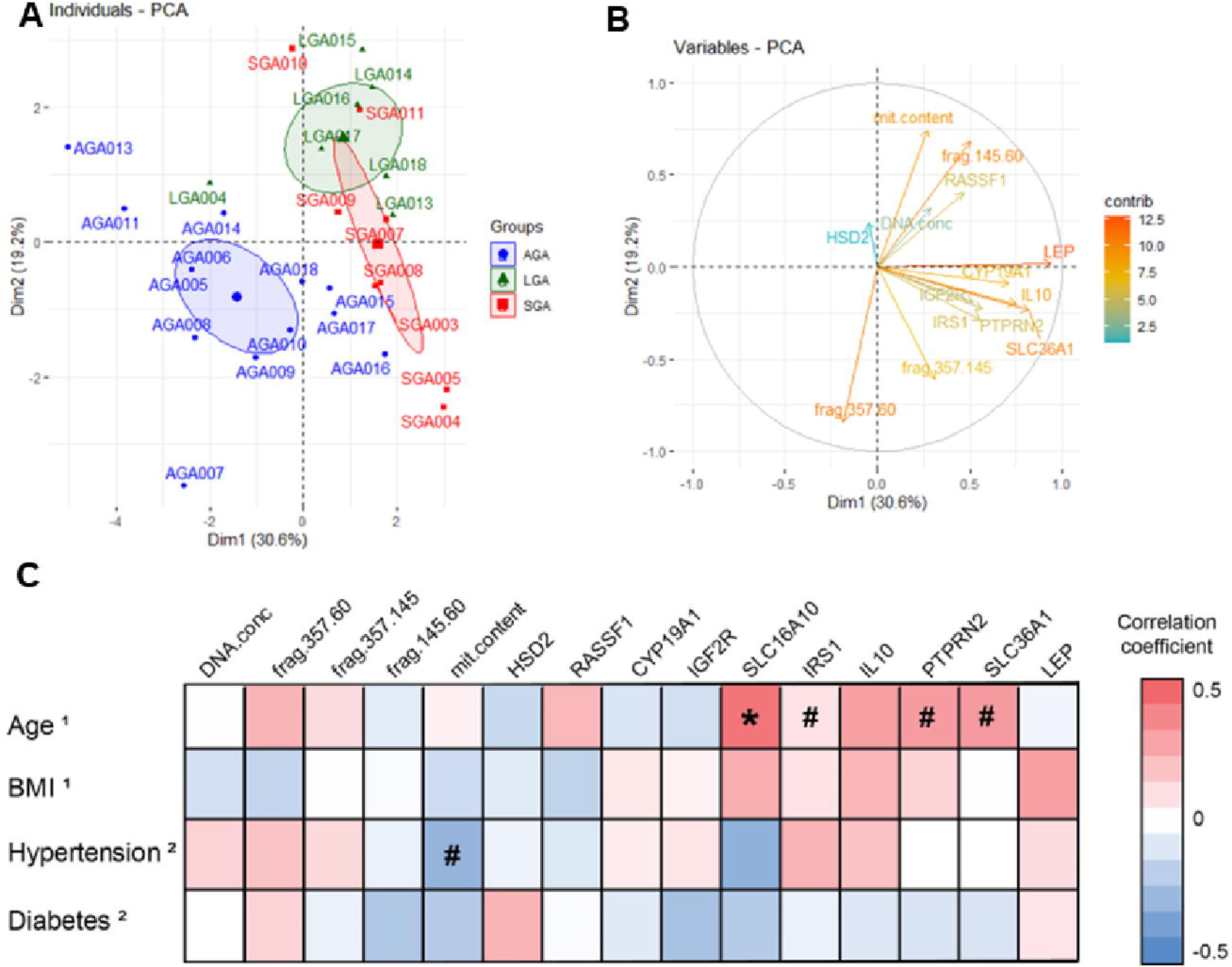
Multivariate analysis in SGA, LGA and AGA pregnancies. A) Samples corresponding to SGA (red), LGA (green) and AGA (blue) pregnancies clustered separately in PCA analysis. SGA, AGA and LGA groups are represented by red, blue and green dots and elipses, respectively. B) Graphic of variables depicting the contribution of each variable in sample distribution. Positively and negatively correlated variables point to the same or opposite side of the plot, respectively. The contributions to the sample discrimination are color coded in a gradient from red (higher contribution) over yellow to light blue (lower contribution). C) Correlation matrix of cirDNA profiles with clinical variables. Correlation coefficients (r) are represented in a color scale from 0.5 (red) to -0.5 (blue). Correlations were calculated using Pearson (^1^) or Biserial (^2^) tests. *: p<0.05, #: p<0.1.

### Building and testing a molecular signature for non-invasive early diagnosis of FGD

Whether the variables assessed in cirDNA samples, can be combined as a marker panel for early detection of FGD pregnancies (Figure 4) was explored using generalized logistic models (GLM) (Supplementary Table S2). Such models aimed at distinguishing between AGA and LGA/SGA pregnancies. The performance of each model was evaluated using three statistical methods (i.e., empirical, binomial, and non-parametric) and a cross-validation strategy. The first model (Figure 4A) has a high power (86.9% accuracy, 80.0% sensitivity and 92.3% specificity) to discriminate FGDs in general, including both SGA and LGA, from AGA pregnancies (AUCs=0.96, 0.98, and 0.96, for empirical, binomial, and non-parametric ROCs, respectively). The performance of the model was further evaluated according to positive predictive and negative predictive values for FGDs (PPV and NPV, respectively), achieving 88.8% PPV and 85.7% NPV. A second model (Figure 4B) has high power (87.5% accuracy, 75.0% sensitivity and 91.7 % specificity) to differentiate specifically SGA pregnancies (AUCs=0.95, 0.96, and 0.93 for empirical, binomial, and non-parametric ROCs, respectively; PPV=75.0% and 91.7 % NPV). The third model (Figure 4C) has high power (93.3% accuracy, 66.7% sensitivity, and 100.0 % specificity) to differentiate specifically LGA pregnancies (AUCs=0.97, 0.98, and 0.96 for empirical, binomial, and non-parametric ROCs, respectively; PPV=100.0% and 92.3 % NPV). Lastly, we generated a fourth model (Figure 4D) with high power (81.2% accuracy, 75.0 % sensitivity, and 83.3 % specificity) to differentiate between SGA and LGA pregnancies (AUCs=0.88, 0.92, and 0.87 for empirical, binomial, and non-parametric ROCs, respectively; PPV=60.0% and 90.9 % NPV).

**Figure 4:**
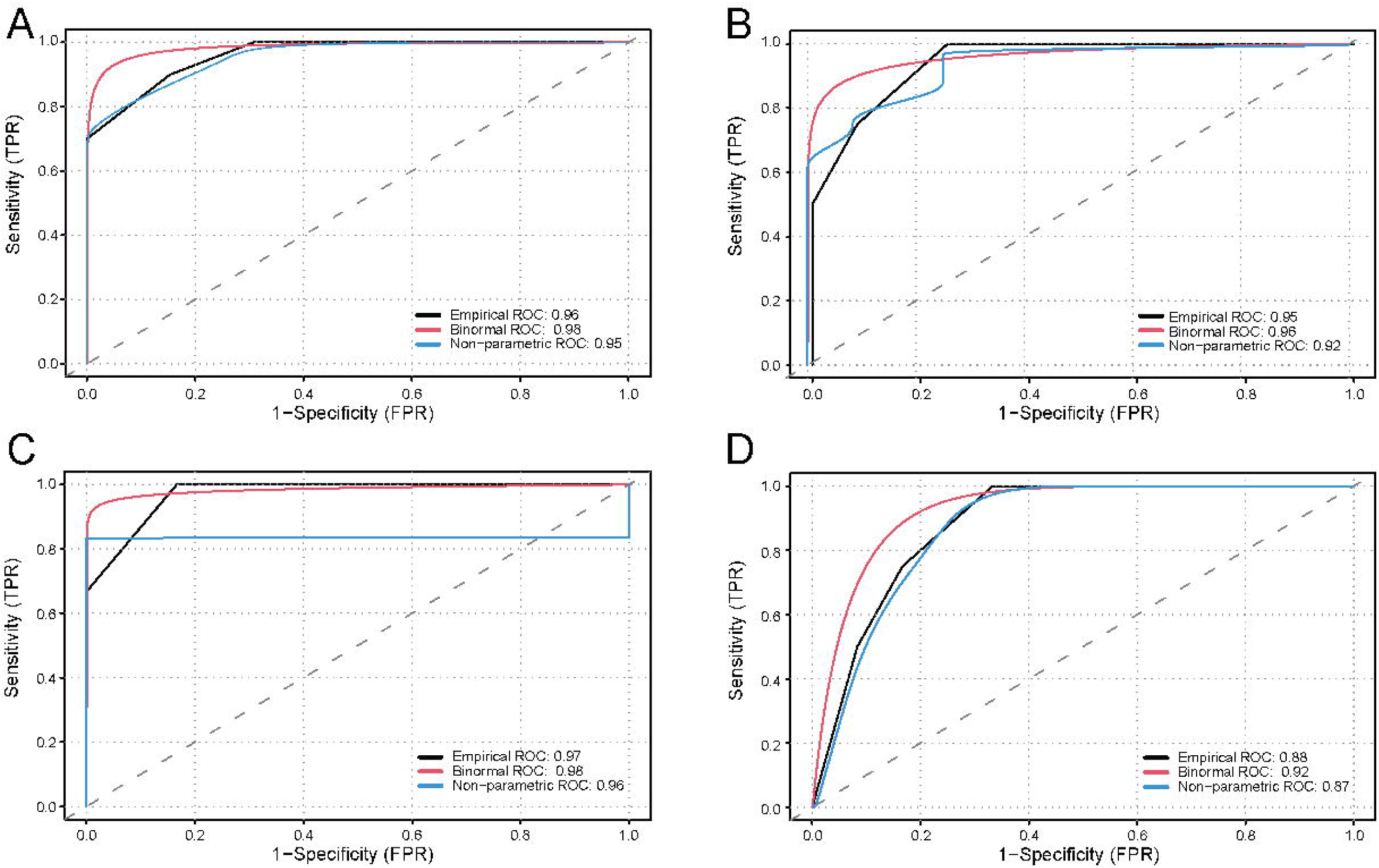
Receiving-Operating Curve (ROC) analysis assessing the performance of the molecular signature for the discrimination between SGA, LGA and AGA pregnancies. The performance of the signature was assessed by calculating the Area Under the Curve (AUC) when applying three different models: Empirical, Binormal and Non-Parametric, represented as black, red, and blue lines, respectively. A) ROC analysis assessing the performance of the molecular signature for the discrimination between AGA and SGA/LGA pregnancies. AUC=0.96, 0.98, and 0.95 for the empirical, binormal, and non-parametric modelling, respectively. B) ROC analysis assessing the performance of the molecular signature for the discrimination between AGA and SGA pregnancies. AUC=0.95, 0.96, and 0.92 for the empirical, binormal, and non-parametric modelling, respectively. C) ROC analysis assessing the performance of the molecular signature for the discrimination between AGA and LGA pregnancies. AUC=0.97, 0.98, and 0.96 for the empirical, binormal, and non-parametric modelling, respectively. D) ROC analysis assessing the performance of the molecular signature for the discrimination between SGA and LGA pregnancies. AUC=0.88, 0.92, and 0.87 for the empirical, binormal, and non-parametric modelling, respectively.

## DISCUSSION

Using a comprehensive cirDNA profiling of maternal blood samples, we report here a novel marker panel enabling accurate prediction of FGDs that can be implemented as early as the first gestational trimester. Our marker panel combines the total amount of cirDNA, with physical characterization of the cirDNA (i.e., fragmentation analysis and ratio of mitochondrial DNA) which are representative of the increase shedding of cirDNA into circulation upon cellular turnover. Furthermore, the proposed panel includes DNA methylation assessment of 10 genes that were selected *a priori* due to their direct roles in fetal growth and placental homeostasis.

Analyses of circulating nucleic acids (i.e., DNA and RNA) in maternal blood are widely used as a non-invasive prenatal diagnostic test (NIPT). Numerous tests and applications have been developed during the last two decades demonstrating the utility and precision of such tests for determining fetal health (e.g., chromosomal alterations), as well as for predicting obstetric complications, such as pre-eclampsia and premature birth. Nevertheless, some controversy remains regarding the clinical utility of cirDNA testing during pregnancy based on the limited fraction of fetal DNA in the total circulating DNA of maternal blood, especially in early gestation (23). Hence, despite the tremendous advances in the field, the use of liquid biopsies for monitoring fetal growth and FGD detection is currently unavailable. A recent study including over 16,000 individuals(24), demonstrated that the amount of cirDNA in maternal blood shows a steady increase during pregnancy with inflection points at the 10th, 19th and 30th week of gestation. Importantly, this study demonstrated that the concentration of cirDNA in maternal blood reached the standard NIPT requirements already at the 9^th^ week of gestation(24), encouraging the interrogation of early fetal markers using high sensitivity approaches.

Large-scale DNA methylation profiles have been proven as a successful source for candidate biomarkers, but their application as molecular diagnostics is hampered by costs and other intrinsic limitations (e.g., the generation of information that is beyond the clinical application). Hence, clinical assay design requires the reduction of the number of measured loci to a manageable level. In this study, we reported a 15-marker signature that can differentiate with high precision (> 90.0% accuracy) between FGD and normal pregnancies in first trimester maternal blood samples. Noteworthy, we did not detect significant differences in the maternal weight, height and BMI at baseline among the SGA, AGA and LGA groups, nor the markers quantification correlated with maternal phenotype. These findings suggest that the molecular signature is not biased towards the identification of constitutionally large or small babies, but rather identify clinically relevant FGDs. As the number of patients per group in this study does not provide enough statistical power to further stratify the SGA and LGA groups according to maternal baseline assessment, further investigation of the performance of the molecular signature in constitutionally large or small babies is warranted.

The performance of the markers discovered in the present study and their use in a clinical setting will be formally assessed in future studies (i.e., assay development, verification, and validation) for single or multi-marker panels using larger datasets. The clinical utility of such marker panels will be further enhanced by combining them with clinical and demographic information (e.g., BMI, maternal and gestational age, diabetes, hypertension, etc.) using multivariate models towards the operationalization of personalized diagnostic and monitoring algorithms in FGD. The application of such algorithms in a clinical setting holds the potential to enable a disruptive path toward precision medicine in FGD.

The presence of fetal DNA in maternal plasma has been reported almost 30 years ago by Dennis Lo and cols(9). Since then, numerous studies have been conducted to determine the origin and dynamics of the fetal cirDNA in maternal plasma, reviewed in (10). cirDNA is shed into maternal blood via apoptosis, necrosis, and oxidative stress(11). These physical characteristics are faithfully represented in the marker panel by inclusion of the cirDNA fragmentation and mitochondrial/nuclear cirDNA ratio. Experimental evidence indicates that cirDNA in plasma derives from the fetus, the placenta and the mother, and there is a dynamic on the relative contribution of each source to the total amount of DNA recovered from maternal plasma or serum, with a rapid clearance after birth (12). Thus, the assessment of cirDNA in maternal blood can therefore provide valuable evidence for fetoplacental health and disease throughout pregnancy (10).

Since epigenetic alterations represent a common trait in most complex diseases, they hold great potential as robust and precise biomarkers in tissues and bodily fluids(25). In particular, the analysis of cirDNA in maternal blood is of outstanding clinical relevance and it is currently a standard practice in NIPT. Several features of DNA methylation support their potential as early diagnostic biomarkers, including the large number of DNA methylation changes in affected cells, their early occurrence in disease development, and their stability enabling detection in small amounts of DNA. DNA methylation is *de facto* being already used in clinical practice(25). DNA methylation biomarkers are instrumental in oncology and several commercial tests based on DNA methylation markers are already implemented in the clinics.

This study provided empirical evidence demonstrating that cirDNA isolated from maternal blood samples at the first gestational trimester has a diagnostic value for the occurrence of FGD. By combining the cirDNA quantification with physical and epigenetic markers, the proposed molecular signature provides a comprehensive profiling of cirDNA in maternal blood. The relatively low number of markers in the panel (i.e., 15 markers) would enable the development of molecular diagnostic assays for early FGD diagnostics that can be implemented in the clinical practice.

Notwithstanding the high potential for the findings, the molecular signature should be further validated using larger independent larger datasets. We anticipate that this signature will be the foundation for the development FGD molecular tests following the Design Control Guidelines for assay development, analytical verification, and clinical validation(26) in future studies. Moreover, the development of diagnostic algorithms combining molecular and clinical data will require a larger number of subjects per group than those in the present study.

In conclusion, our findings show that maternal blood cirDNA profiles accurately detects early gestation FGD. The proposed novel marker panel hold great potential for implementation of low invasive approaches for reliable prediction of FGDs as early as the first trimester of pregnancy.

## METHODS

### Subject recruitment

Sex as a biological variable was not considered in this study. Pregnant participants were recruited for this study. Figure 1 summarizes the study design. Pregnant subjects were prospectively recruited at their initial Obstetrics visit, blood samples were collected by a certified phlebotomist during their first 9-13 week of gestation and plasma fractions were separated as detailed below. Maternal demographic and clinical information (Table 1), as well as ultrasound data from prenatal controls was prospectively retrieved from existing electronic medical records. After birth and based on the outcome of the pregnancy, subjects were separated into three groups SGA (n=11), LGA (n=18) and AGA (n=29). SGA and LGA pregnancies were defined taking in account gestational age and weight at delivery, following the revised reference chart for the US(27). Pregnancies with babies above 90% and below 10% percentiles were regarded as LGA and SGA, respectively. No significant differences (p>0.05) were detected in maternal age, ethnicity, height, weight and BMI at NOB among the subjects on the SGA, AGA and LGA groups (Table 1).

**Table 1:** Subjects demographics

| Group | SGA | AGA | LGA | p-value |
| --- | --- | --- | --- | --- |
| n | 11 | 29 | 18 |  |
| Maternal age | 28.12 ± 7.71 | 29.6 ± 4.51 | 28.46 ± 4.39 | 0.709<sup>a</sup> |
| [years ± SD] |  |  |  |  |
| Maternal ethnicity | 9 (81.8%) | 28 (96.5%) | 18 (100%) | 0.084<sup>b</sup> |
| [White (%)] |  |  |  |  |
| Maternal height | 1.59 ± 6.23 | 1.64 ± 8.44 | 1.63 ± 5.41 | 0.108<sup>a</sup> |
| [meters ± SD] |  |  |  |  |
| Maternal weight | 75.78 ± 12.22 | 78.42 ± 19.91 | 85.35 ± 26.24 | 0.716<sup>a</sup> |
| [kilograms ± SD] |  |  |  |  |
| Maternal BMI at NOB | 30.0 ± 3.86 | 29.0 ± 7.14 | 31.2 ± 9.83 | 0.547<sup>a</sup> |
| [kg/m2 ± SD] |  |  |  |  |
| Gestational Age at | 37.9 ± 1.64 | 38.8 ± 1.12 | 39.2 ± 1.11 | 0.078<sup>a</sup> |
| Birth [weeks ± SD] |  |  |  |  |
| Birth weight | 2433.32 ± 506.63 | 3219.91 ± 340.46 | 4030.54 ± 221.23 | <0.001<sup>a</sup> |
| [g ± SD] |  |  |  |  |

<sup>a</sup>: Kruskal-Wallis test
<sup>b</sup>: $\chi^2$ test, white vs. non-white

### Plasma cirDNA isolation from whole blood maternal samples

Plasma and cellular fraction from whole blood samples was separated by centrifugation at 1,800 rpm for 20 minutes. cirDNA from the plasma fraction was isolated using the Circulating Nucleic Acids kit (Qiagen, Valencia, CA), according to manufacturer’s instructions. Isolated cirDNA was stored at -80°C until use.

### Plasma cirDNA characterization

cirDNA amounts and fragmentation for each sample was quantitively assessed using SYBR-green based qPCR assays, as previously reported (28). In brief, total cirDNA amount was assessed by single-locus (i.e., KRAS) amplification and extrapolation to calibration curves. cirDNA fragmentation was assessed using a combination of three qPCR assays targeted to the same locus (i.e., KRAS). These assays share the same reverse primer, but the length of the assay differs due to the forward primer expanding to i) 357 bp representing long DNA fragments, ii) 145 bp representing nucleosome-sized DNA fragments, and iii) 60 bp representing highly fragmented DNA. The relative enrichment of these three fractions was calculated as the ratio between the amplification (i.e., DDCt) and the DNA Fragmentation Index (DFI) was calculated for each sample. Mitochondrial DNA release into maternal blood was quantified by the Mitochondrial-Nuclear DNA ratio (MNR) according to the methods of Otandault, et. al(29). cirDNA methylation was measured using Methylation-Sensitive Restriction Enzyme qPCR (MSRE-qPCR), as previously reported (30), and encompassed 10 candidate genes selected according to previous reports on their involvement in placental homeostasis and fetal development (*HSD2, RASSF1, CYP19A1, IGF2R, SLC16A10, IRS1, IL10, PTPRN2, SLC36A1, LEP*) (13–22). Primers for all qPCR assays are provided in Supplementary Table S1.

### Building of molecular signature

All data analysis was conducted in R (version 4.2.2). Principal component analysis and variable plots were conducted using the *factoextra* package version 1.0.7. The molecular signature was extracted based on cirDNA markers and clinical attributes, and combined in generalized logistic models (GLM), according to the following comparisons: i) AGA vs. LGA/SGA pregnancies, ii) AGA vs. SGA pregnancies, iii) AGA vs. LGA pregnancies, and iv) SGA vs. LGA pregnancies (Supplementary Table S2). GLMs were build using the *riskRegression* (version 1.3.7) package. Empirical, binormal, and non-parametric statistical methods, each using *randomForest* (version 4.7.1), were the parameters leveraged in the construction of three ROC curves using R-package *ROCit* (version 2.1.1).

### Statistics

Univariate data analysis and plotting were conducted using GraphPad Prism 10 (version 10.1.2; GraphPad Software, Boston, MA). Differences in continuous clinical variables and cirDNA quantity, fragmentation, mitochondrial DNA content and methylation between AGA, SGA and LGA were assessed using Kruskal-Wallis test, followed by multiple test comparisons using Dunn’s test implemented in the R software (version 4.4.2). Differences in categorical clinical variables were assessed using the χ^2^ test implemented in the R software (version 4.4.2). Principal component analysis was conducted using the R-package *factoextra* (version 1.0.7). Correlations were calculated using Pearson or Biserial methods implemented in the R software (version 4.4.2). Molecular signatures were built and tested as described above.

### Study approval

This project has been evaluated and approved by the Institutional Review Board (IRB) of the University of Missouri (protocol number: 2016992_MU) and it was conducted following the ethical standards for human experimentation established in the Declaration of Helsinki. A written informed consent was received from each participant prior to participation.

### Data sharing

To ensure independent interpretation of clinical study results and enable authors to fulfill their role and obligations under the International Committee of Medical Journal Editors (ICMJE) criteria, the authors grant all external authors access to data pertinent to the development of the publication. Scientific and medical researchers can access to molecular and anonymized clinical data and supporting analytic code utilized in this publication by contacting the corresponding author.

## AUTHOR’S CONTRIBUTION

R.C. participated in the conceptual framework of the project, performed experiments, analyzed data, and wrote the manuscript. K.C., G.S., M.O. and M.R. performed experiments. J.H. conducted data analysis. H.W., D.G. and J.R.G provided medical expert guidance, participated in data analysis and interpretation of results, and served as blinded observers. All authors have reviewed and approved the final version of the manuscript.

## Data Availability

All data produced in the present study are available upon reasonable request to the authors

## ACKNOWLEDGMENTS

Authors would like to acknowledge the Research Success Center (RSC) of the Department of Obstetrics, Gynecology & Women’s Health at the University of Missouri for patient recruitment and sample collection.

Parts of this study were presented at the 90^th^ Annual Meeting of the Central Association of Obstetricians and Gynecologists. Nashville TN. USA. October 26-28, 2023.

## CONFLICT OF INTERESTS

The authors report no conflict of interest.

## TRIAL REGISTRATION

Not applicable

## FUNDING

Internal startup funds from the University of Missouri to RC. The funding source was not involved in the study design; in the collection, analysis and interpretation of data; in the writing of the report; and in the decision to submit the article for publication.

## SUPPLEMENTARY MATERIAL

**Supplementary Table S1:** qPCR primers for cirDNA characterization.

**Supplementary Table S2:** Generalized logistic models for AGA, SGA and LGA molecular signatures.

## Notes

### Competing Interest Statement

The authors have declared no competing interest.

### Funding Statement

This study did not receive any funding

