## Supplemental Material for "Fetal Growth Disorders Detection During First Trimester Gestation Through Comprehensive Maternal Circulating DNA Profiling"

**Contents:**

1. Supplementary Table S1: qPCR primers for cirDNA characterization.
2. Supplementary Table S2: Generalized logistic models for AGA, SGA and LGA molecular signatures.

**Supplementary Table S1:** qPCR primers for cirDNA characterization.

| *1. DNA quantification primers* | | | | |
|---|---|---|---|---|
| **Locus name** | **Fragment Size** | **F primer sequence** | **R primer sequence** | |
| **KRAS** | 189 | aatccgtgtgggtcagagag | gaaacaatagccaccctcctt | |
| *2. DNA fragmentation primers* | | | | |
| **Locus name** | **Fragment Size** | **F primer sequence** | **R primer sequence** | |
| **KRAS** | 60 | GCCTGTGACCTACAGTGAAAA | TGACCAAGCAAAACAGACCA | |
| **KRAS** | 145 | TGGGCTGTGACATTGCTG | TGACCAAGCAAAACAGACCA | |
| **KRAS** | 357 | GGCATCTCTAGGACGAAGGT | TGACCAAGCAAAACAGACCA | |
| *3. Mitochondrial DNA quantification primers* | | | | |
| **Locus name** | **Fragment Size** | **F primer sequence** | **R primer sequence** | |
| **COX3_Mit** | 61 | CTCCTACAAGCCTCAGAGTAC | GAGCCGTAGATGCCGTCG | |
| **COX3_Mit** | 253 | TTAGGAGGGCACTGGCCC | GAGCCGTAGATGCCGTCG | |
| *4. qMSRE-Primers* | | | | |
| **Locus name** | **Fragment Size** | **F primer sequence** | **R primer sequence** | **Restriction enzyme** |
| **HSD2** | 132 | AGGGGTGAGCGCGCCTTA | AGGAACCAGCCGGTGCTC | *BstUI* |
| **RASSF1** | 115 | ATGCGCAGCGCGTTGGCA | CCCAACCGGGCCATGTC | *BstUI* |
| **CYP19A1** | 132 | TTGGCTGTACGCCGAGGTC | CACCCCGGTCGCGTCTC | *BstUI* |
| **IGF2R** | 112 | AGGCCGCGGTTCTCTGC | TGCATCCCTCGCCGGTAG | *BstUI* |
| **SLC16A10** | 115 | GCTGGTAACTCGCGTCCCT | AGCGGCTGCGCCTCGCT | *BstUI* |
| **IRS1** | 129 | GTTGATGTTGAAGCAGCTCTCA | GTACTGCGCGCGGCCAG | *BstUI* |
| **IL10** | 108 | ACCATGTTGACCAGGCTGGTTA | TTCTAGGCCGGGCGCGGT | *BstUI* |
| **PTPRN2** | 116 | AGCCGGACGCGGTGGCT | GAGATGGAGTTTCACTATATTGCT | *BstUI* |
| **SLC36A1** | 120 | AGTGCAGTGGCGCGATCT | AAATTAGCCGGGCGTGGTA | *BstUI* |
| **LEP** | 150 | AAGGGTGCGCGCGTGGC | GCAGGCCCCGGCGCATT | *BstUI* |

**Supplementary Table S2:** Generalized logistic models for AGA, SGA and LGA molecular signatures.

| *AGA compared with SGA and LGA pregnancies combined* | |
|---|---|
| **Variable** | **Coefficient** |
| (Intercept) | 4.385498009 |
| DNA.conc | -0.01279025 |
| frag.357.60 | -2.44131494 |
| frag.357.145 | 1.24497366 |
| frag.145.60 | -1.60747438 |
| HSD2.methylation | -2.89507858 |
| RASSF1.methylation | 0.146718432 |
| CYP19A1.methylation | 11.58241807 |
| IGF2R.methylation | 0.585137948 |
| SLC16A10.methylation | -2.153820021 |
| IRS1.methylation | -5.270003935 |
| IL10.methylation | -10.51680143 |
| PTPRN2.methylation | 9.572904547 |
| SLC36A1.methylation | 11.94714501 |
| LEP.methylation | -4.792703454 |

| *AGA compared with SGA pregnancies* | |
|---|---|
| **Variable** | **Coefficient** |
| (Intercept) | -393.9689287 |
| DNA.conc | -4.139110129 |
| frag.357.60 | 39.31930104 |
| frag.357.145 | -11.69993123 |
| frag.145.60 | 165.5102135 |
| HSD2.methylation | 212.466937 |
| RASSF1.methylation | 768.4719706 |
| CYP19A1.methylation | -1349.656813 |
| IGF2R.methylation | -68.14517047 |
| SLC16A10.methylation | 267.6507411 |
| IRS1.methylation | 46.95964117 |
| IL10.methylation | -173.7575997 |
| PTPRN2.methylation | -152.2931863 |
| SLC36A1.methylation | 88.03329019 |
| LEP.methylation | -146.9948932 |

| *AGA compared with LGA pregnancies* | |
|---|---|
| **Variable** | **Coefficient** |
| (Intercept) | -511.6783041 |
| DNA.conc | -0.943862588 |
| frag.357.60 | 46.96788678 |
| frag.357.145 | -9.292666181 |
| frag.145.60 | 140.8999876 |
| HSD2.methylation | 116.9013711 |
| RASSF1.methylation | 12.17467052 |
| CYP19A1.methylation | -1.23183611 |
| IGF2R.methylation | -92.58511867 |
| SLC16A10.methylation | 46.36299955 |

| | |
|---|---|
| IRS1.methylation | 67.56356887 |
| IL10.methylation | -123.1021541 |
| PTPRN2.methylation | 17.62002955 |
| SLC36A1.methylation | 168.2267322 |
| LEP.methylation | -15.45504754 |

***SGA compared with LGA pregnancies***

| **Variable** | **Coefficient** |
|---|---|
| (Intercept) | -393.9689287 |
| DNA.conc | -4.139110129 |
| frag.357.60 | 39.31930104 |
| frag.357.145 | -11.69993123 |
| frag.145.60 | 165.5102135 |
| HSD2.methylation | 212.466937 |
| RASSF1.methylation | 768.4719706 |
| CYP19A1.methylation | -1349.656813 |
| IGF2R.methylation | -68.14517047 |
| SLC16A10.methylation | 267.6507411 |
| IRS1.methylation | 46.95964117 |
| IL10.methylation | -173.7575997 |
| PTPRN2.methylation | -152.2931863 |
| SLC36A1.methylation | 88.03329019 |
| LEP.methylation | -146.9948932 |
